# Evaluating the Performance of Large Language Models on a Neurology Board-Style Examination

**DOI:** 10.1101/2023.07.13.23292598

**Authors:** Marc Cicero Schubert, Wolfgang Wick, Varun Venkataramani

## Abstract

**Background and Objectives:** Recent advancements in large language models (LLMs) such as GPT-3.5 and GPT-4 have shown impressive potential in a wide array of applications, including healthcare. While GPT-3.5 and GPT-4 showed heterogeneous results across specialized medical board examinations, the performance of these models in neurology board exams remains unexplored.

**Methods:** An exploratory, prospective study was conducted between May 17 and May 31, 2023. The evaluation utilized a question bank approved by the American Board of Psychiatry and Neurology, designed as part of a self-assessment program. Questions were presented in a single best answer, multiple-choice format. The results from the question bank were validated with a small question cohort by the European Board for Neurology. All questions were categorized into lower-order (recall, understanding) and higher-order (apply, analyze, synthesize) questions. The performance of GPT-3.5 and GPT-4 was assessed in relation to overall performance, question type, and topic. In addition, the confidence level in responses and the reproducibility of correctly and incorrectly answered questions was evaluated. Univariable analysis was carried out. Chi-squared test and Bonferroni correction were used to determine performance differences based on question characteristics. To differentiate characteristics of correctly and incorrectly answered questions, a high-dimensional tSNE analysis of the question representations was performed.

**Results:** In May 2023, GPT-3.5 correctly answered 66.8 % of 1956 questions, whereas GPT-4 demonstrated a higher performance level, correctly answering 85 % of questions in congruence with near-passing and passing of the neurology board exam. GPT-4’s performance surpassed both GPT-3.5 and question bank users (mean human user score: 73.8%). An analysis of twenty-six question categories showed that GPT-4 outperformed human users in Behavioral, Cognitive and Psych-related questions and demonstrated superior performance to GPT-3.5 in six categories. Both models performed better on lower-order than higher-order questions according to Bloom Taxonomy for learning and assessment (GPT4: 790 of 893 (88.5%) vs. 872 of 1063 (82%), GPT-3.5: 639 of 893 (71.6%) vs. 667 of 1063 (62.7%)) with GPT-4 also excelling in both lower-order and higher-order questions. The use of confident language was observed consistently across both models, even when incorrect (GPT-4: 99.3%, 292 of 294 incorrect answers, GPT-3.5: 100%, 650 of 650 incorrect answers). Reproducible answers of GPT-3.5 and GPT-4 (defined as more than 75 % same output across 50 independent queries) were associated with a higher percentage of correct answers (GPT-3.5: 66 of 88 (75%), GPT-4: 78 of 96 (81.3%)) than inconsistent answers, (GPT-3.5: 5 of 13 (38.5%), GPT-4: 1 of 4 (25%)). Lastly, the high-dimensional embedding analysis of correctly and incorrectly answered questions revealed no clear differentiation into distinct clusters.

**Discussion:** Despite the absence of neurology-specific training, GPT-4 demonstrated commendable performance, whereas GPT-3.5 performed slightly below the human average question bank user. Higher-order cognitive tasks proved more challenging for both GPT-4 and GPT-3.5. Notwithstanding, GPT-4’s performance was equivalent to a passing grade for specialized neurology board exams. These findings suggest that with further refinements, LLMs like GPT-4 could play a pivotal role in applications for clinical neurology and healthcare in general.

## Introduction

Deep learning algorithms have been investigated in neurology for a variety of tasks, such as neurologic diagnosis, prognosis and treatment ^1,2^. However, the role and potential application of large language models (LLMs) in neurology have been unexplored. The recent emergence of the powerful transformer-based AI models GPT-3.5 and GPT-4 ^3,4^ provides a new avenue for exploring their implications in the field of neurology. These Large Language Models (LLMs) undergo training using expansive datasets, encompassing more than 45 terabytes of information. This rigorous training process equips them to recognize patterns and associations among words, which, in turn, empowers them to produce responses that are both contextually accurate and logically consistent ^5^. The application of these models in specialized medical examinations has been tested to some extent. GPT-3.5 showed near-pass performance in the United States Medical Licensing Examination (USMLE) ^6,7^ while it failed to pass the ophthalmology board examination ^8^. Two recent reports showed slightly deviating results on neurosurgery board-style exams with one report claiming a near-pass with GPT-3.5 while the other showed an approximately 10 % lower performance. GPT-3.5 achieved a near-pass in radiology board-like examinations while GPT-4 in neurosurgery board-like examinations successfully passed it ^9^. In contrast, neurology board-like exams present a different set of challenges. As compared to radiology, ophthalmology, or neurosurgery, the questions in neurology board examinations often present complex narratives with subtle diagnostic clues that require a nuanced understanding of neuroanatomy, neuropathology, and neurophysiology. The candidate is expected to navigate through these complex narratives, extracting relevant data, and synthesizing this information into a coherent diagnostic hypothesis and subsequent therapeutic decisions. Written board examinations, designed to test a broad range of neurology topics, are common in the US, Canada and Europe. These examinations typically employ multiple-choice questions, a format also adopted in the US by the American Board of Psychiatry and Neurology ^10^, and in Europe by the European Board of Neurology (UEMS-EBN) ^11^.

In this exploratory study, our objective was to evaluate the performance of GPT-3.5 and GPT-4 in comparison to human performance in neurology board-like written examinations. We used the context of neurology board-like written examinations as a representative example to scrutinize the complex reasoning abilities and the capacity of large language models (LLMs), specifically GPT-3.5 and GPT-4, to navigate intricate medical cases, thereby illuminating their potential in more sophisticated, real-world clinical applications. Our ultimate aim was not only to determine their accuracy and reliability in this specialized context but also to characterize their strengths and limitations. As LLMs continue to evolve, understanding their potential contributions and challenges in medical examinations could pave the way for future applications in neurology and neurology education.

## Methods

This exploratory prospective study was performed from May 17 to May 31, 2023.

### Standard Protocol Approvals, Registrations, and Patient Consents

This study did not involve human subjects or patient data, so it was exempt from institutional review board approval.

### Multiple-Choice Question Selection and Classification

A question bank resembling neurology board questions consisting of 2036 questions (from boardvitals.com) ^12^ was used for the evaluation of GPT-3.5 and GPT-4. Questions including videos or images as well as questions that were based on preceding questions were excluded in this study (n=80 questions excluded, n=1956 questions included). This question bank is approved by the American Board of Psychiatry and Neurology (ABPN) as part of a Self-Assessment program and can be used as a tool for certified medical education ^12^. Questions were in single best answer, multiple-choice format with three, four or five distractors and one correct answer. To validate the results from this question bank, open-book sample questions from 2022 from the European Board of Neurology were used (n=19 questions). These questions are either behind a paywall (in the case of the question bank) or published after 2021 and therefore out-of-training data for both GPT-3.5 and GPT-4.

Questions were then classified by type using principles of Bloom Taxonomy for learning and assessment as testing either lower-order (remembering, basic understanding) or higher-order (applying, analyzing, or evaluating) thinking ^13,14^. We both let GPT-4 and the investigators evaluate whether the questions were in the lower-order or higher-order category and the investigators discussed cases of incongruencies. GPT-4 classified in accordance with the investigators in 87.5% (175 of 200 questions), GPT-3.5 in 84.5% (169 of 200 questions).

The questions can be further categorized according to 26 topics in the field of neurology that are listed in Table 1. Performance by users per individual question was available from the test portal while this information was not available for the sample questions from the EBN.

**Table 1:** Performance of GPT-3.5, GPT-4 and Question Bank Users by Question Type and Topic.

| Question Type | Question N | Human Correct Mean % | GPT-3.5 Correct N (%) | GPT-4 Correct N (%) | Adj. P Value GPT-3.5 vs Human | Adj. P Value GPT-4 vs Human | Adj. P Value GPT-3.5 vs GPT-4 |
| --- | --- | --- | --- | --- | --- | --- | --- |
| All Questions | 1956 | 73.8 | 1306 (66.8) | 1662 (85) | <0.0001 | <0.0001 | <0.0001 |
| Order of thinking |  |  |  |  |  |  |  |
| Higher | 1063 | 73.9 | 667 (62.7) | 872 (82) | <0.0001 | <0.0001 | <0.0001 |
| Lower | 893 | 73.6 | 639 (71.6) | 790 (88.5) | 0.73 | <0.0001 | <0.0001 |
| Category |  |  |  |  |  |  |  |
| Basic Neuroscience | 128 | 74.1 | 83 (64.8) | 109 (85.2) | 1 | 1 | 0.008 |
| Behavioral, Cognitive, Psych | 482 | 76 | 362 (75.1) | 433 (89.8) | 1 | <0.0001 | <0.0001 |
| Cerebrovascular | 113 | 77.7 | 77 (68.1) | 93 (82.3) | 1 | 1 | 0.54 |
| Child Neurology | 73 | 69.8 | 48 (65.8) | 57 (78.1) | 1 | 1 | 1 |
| Congenital | 20 | 74.3 | 14 (70) | 18 (90) | 1 | 1 | 1 |
| Cranial Nerves | 46 | 70 | 24 (52.2) | 34 (73.9) | 1 | 1 | 1 |
| Critical Care | 28 | 71.3 | 15 (53.6) | 25 (89.3) | 1 | 1 | 0.2 |
| Demyelinating Disorders | 49 | 81.9 | 37 (75.5) | 45 (91.8) | 1 | 1 | 1 |
| Epilepsy, Seizures | 55 | 73.7 | 34 (61.8) | 39 (70.9) | 1 | 1 | 1 |
| Ethics | 5 | 84.8 | 2 (40) | 4 (80) | 1 | 1 | 1 |
| Genetic | 20 | 70.4 | 16 (80) | 17 (85) | 1 | 1 | 1 |
| Headache | 59 | 74 | 40 (67.8) | 50 (84.7) | 1 | 1 | 1 |
| Imaging/Diagnostic Studies | 10 | 65.4 | 5 (50) | 9 (90) | 1 | 1 | 1 |
| Movement Disorders | 91 | 75.1 | 54 (59.3) | 79 (86.8) | 1 | 1 | 0.002 |
| Neuro-Ophthalmology | 46 | 72.4 | 29 (63) | 40 (87) | 1 | 1 | 0.42 |
| Neuro-Otology | 42 | 66.9 | 20 (47.6) | 31 (73.8) | 1 | 1 | 0.66 |
| Neuroinfectious Disease | 30 | 68.1 | 16 (53.3) | 25 (83.3) | 1 | 1 | 0.69 |
| Neurologic Complications of Systemic Disease | 42 | 71.8 | 21 (50) | 31 (73.8) | 1 | 1 | 1 |
| Neuromuscular | 184 | 68.9 | 120 (65.2) | 145 (78.8) | 1 | 1 | 0.14 |
| Neurotoxicology, Nutrition, Metabolic | 93 | 70.4 | 63 (67.7) | 82 (88.2) | 1 | 0.1 | 0.04 |
| Oncology | 72 | 70 | 41 (56.9) | 62 (86.1) | 1 | 0.71 | 0.006 |
| Pain | 65 | 78.8 | 42 (64.6) | 58 (89.2) | 1 | 1 | 0.05 |
| Pharmacology | 72 | 74 | 48 (66.7) | 60 (83.3) | 1 | 1 | 0.89 |
| Pregnancy | 19 | 69.1 | 14 (73.7) | 15 (78.9) | 1 | 1 | 1 |
| Sleep | 92 | 77.4 | 67 (72.8) | 82 (89.1) | 1 | 1 | 0.22 |
| Trauma | 20 | 74.2 | 14 (70) | 19 (95) | 1 | 1 | 1 |

Chi-squared test was used to calculate p-values. P-values were adjusted for multiple testing using the Bonferroni correction.

### Large language models

GPT-3.5 (version: gpt-3.5-turbo (Chat-Completion); OpenAI) and GPT-4 (version: gpt-4 (Chat-Completion), OpenAI) were used via API. These are two commonly used large language models ^3,5^. At the time of this study, we did not have access to other powerful closed-source models such as ClaudeV1 ^15^ or PaLM2 ^16^. GPT-3.5 and GPT-4 have been pretrained on over 45 terabytes of text data, including a substantial portion of internet websites, books, and articles. No additional neurology-specific pretraining was performed. In this study, we used server-contained language models that were trained up to September 2021. The used models do not have the ability to search the internet or external databases.

### Data Collection and Assessment

Each multiple-choice stem along with its answer choices was provided to GPT-3.5 and GPT-4 via its API together with the following prompt:

“You are a medical doctor and are taking the neurology board exam. The board exam consists of multiple choice questions.

All output that you give must be in JSON format.

- Return the answer letter

- Give an explanation

- Rate your own confidence in your answer based on a Likert scale that has the following grades: 1 = no confidence [stating it does not know]; 2 = little confidence [ie, maybe]; 3 = some confidence; 4 = confidence [ie, likely]; 5 = high confidence [stating answer and explanation without doubt])

- Classify the question into the following two categories: 1. lower order questions that probe remembering and basic understanding, and 2. higher order question where knowledge needs to be applied, analysis capabilities are examined, or evaluation is needed. (return “Higher” or “Lower”)

- Rate the confidence of your classification into these categories based on the Likert scale that has the following grades1 = no confidence [stating it does not know]; 2 = little confidence [ie, maybe]; 3 = some confidence; 4 = confidence [ie, likely]; 5 = high confidence [stating answer and explanation without doubt])

Your output must look like the following:

{“answerletter”:…,”reasoning”:…,”confidence_answer_likert”:…,”classification”:…,”confidence_classifi cation_likert”:… “

All answer choices and responses were recorded. A passing score was considered 70% or above on this neurology board–style examination without images to approximate the written examination from the ABPN and the European Board of Neurology (EBN). The question bank uses 70 % as passing threshold to gain credits for certified medical education (CME) points. The Royal College examination in Canada considers 70% or above on all written components a passing score. There, questions undergo psychometric validation, with removal of questions found not discriminatory or too difficult, which was not performed. The ABPN and EBN examinations use criterion-referenced scoring, which was not used.

For all analyses except the reproducibility analyses, the questions were answered once. For the reproducibility analyses, 100 questions were answered by GPT-3.5 and GPT-4 with 50 independent queries probing the principle of self-consistency ^17^.

### High-dimensional analysis of question representations by GPT-3.5 and GPT-4

For the high-dimensional analysis of question representations, the embeddings of these questions were analyzed. These numeric vector representations encompass the semantic and contextual essence of the tokens -in this context, the questions - processed by the model ^18^. The source of these embeddings is the model parameters or weights, which are employed to code and decode the texts for input and output. A dimensionality reduction of the embeddings was performed with a tSNE analysis ^19^ and clusters were subsequently examined.

### Data availability statement

No patient data was used in this study. The prompts used in this study are deposited in the Methods section. Software code used for inquiring the Open AI API has been deposited on GitHub: https://github.com/venkataramani-lab/NeurologyBoard_LLM. Questions from the EBN can be accessed at https://www.uems-neuroboard.org/web/images/docs/exa19m/2023/Example-Questions-selection2023.pdf and questions from the question bank can be found on boardvitals.com. Metadata about the questions (e.g. question length, the models’ score or their embeddings) is available on GitHub: https://github.com/venkataramani-lab/NeurologyBoard_LLM.

### Statistical Analyses

First, the overall performance was evaluated. Next, we compared the performance across different types of questions (namely, lower and higher order) using a single-variable analysis approach (employing the Chi-squared test). We also executed a subgroup analysis for various subclasses of higher-order thinking questions and the 26 topics, where we utilized the Chi-squared test for multiple comparisons with a Bonferroni correction. Given that GPT-3.5 and GPT-4 had a probabilistic chance of correctly answering each question, we utilized a guessing correction formula to glean further understanding ^20^: it is computed by subtracting the ratio of the number of incorrect responses to (the total number of choices minus one) from the number of correct responses:

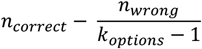

We contrasted the confidence level of responses between correct and incorrect answers by employing the Mann-Whitney U test after testing for normality using the Shapiro-Wilk test. For the correlation analysis between human performance and model performance, human quartiles were converted to numeric values (1-4). A P-value of less than .05 was deemed indicative of a significant difference. All these statistical examinations were carried out in R (version 4.0.5; accessible at https://r-project.org) ^21^.

## Results

### Overall Performance

First, we examined the proficiency of GPT-3.5 and GPT-4 against a question bank set. Evidently, GPT-4 displayed an 85% accuracy level (1662 correct responses out of 1956 questions), superseding GPT-3.5, which managed a 66.8% accuracy level (1306 correct responses out of 1956 questions). When adjusting for random guessing, GPT-4 yielded an 80.9% score (1583 out of 1956), as opposed to GPT-3.5’s 57.8% score (1131 out of 1956). Remarkably, in comparison to the average user of the testing platform (73.8%), GPT-4’s performance was superior (p<0.0001), whereas GPT-3.5 underperformed (p<0.0001, as detailed in Table 1).

To corroborate these results, we also investigated the performance based on openly available sample questions from the EBN for its board examination. Here, GPT-4 correctly responded in 73.7 % of the questions (14 out of 19 questions) while GPT-3.5 only gave a correct response in 52.6% of the questions (10 out of 19 questions), with no significant differences between GPT3.5 and GPT-4 (p=0.31, Supplementary Table 1).

Table 1 shows the performance of GPT 3.5 and 4 as well as the average test user overall and stratified for question type and topic on the user bank while Supplementary Table 1 shows the results on the sample questions of the EBN. Taken together, this demonstrates that GPT-4 is able to pass a neurology board-like exam, whereas GPT-3.5’s performance falls short of passing such a specialized examination.

### Performance By Question Type

Upon analyzing the performance based on question type, it was discernible that both GPT-3.5 and GPT-4 excelled in lower-order questions (GPT-3.5: 639/893 (71.6%), GPT-4: 790/893 (88.5%)) as compared to higher-order questions (GPT-3.5: 667/1063 (62.7%), GPT-4: 872/1063 (82%), p<0.0001, see Table 1).

In the context of lower-order questions, GPT-3.5’s performance (639/893 (71.6%)) was akin to human users’ performance (73.6%, p=0.73). However, GPT-3.5 lagged in answering higher-order questions (667/1063 (62.7%) versus 73.9%, p<0.0001, as exhibited in Table 1)). In both lower and higher-order questions, GPT-4 surpassed GPT-3.5 and human users, marking a great difference (p<0.0001, Table 1).

Supplementary Figures 1-4 provide examples of questions, both correctly and incorrectly answered by GPT-4, categorized into lower and higher-order categories. Interestingly, when segregating the questions into quartiles according to the average performance of human users – easy, intermediate, advanced, and difficult – a correlation became evident between the performance of GPT-3.5, GPT-4, and the average human user (R=0.84, p<0.0001). This correlation potentially suggests shared difficulties faced by humans and these Language Learning Models (LLMs), as depicted in Supplementary Figure 5 and Table 2.

**Table 2:** Comparison of GPT-3.5, GPT-4 and Question Bank Users by Question Type, Difficulty and Topic.

| Order of Thinking | Questions<br>N | Human<br>Correct<br>Mean % | GPT-3.5<br>Correct<br>N (%) | GPT-4<br>Correct<br>N (%) | P Value<br>GPT-3.5<br>vs Human | P Value<br>GPT-4 vs<br>Human | P Value<br>GPT-3.5<br>vs GPT-4 |
| --- | --- | --- | --- | --- | --- | --- | --- |
| **Easy Questions (1<sup>st</sup> quartile)** |  |  |  |  |  |  |  |
| Higher | 283 | 93.3 | 234 (82.7) | 275 (97.2) | 0 | 0.39 | <0.0001 |
| Lower | 226 | 92.9 | 199 (88.1) | 214 (94.7) | 0.87 | 1 | 0.15 |
| **Intermediate Questions (2<sup>nd</sup> quartile)** |  |  |  |  |  |  |  |
| Higher | 271 | 82.1 | 189 (69.7) | 251 (92.6) | 0.01 | 0.002 | <0.0001 |
| Lower | 237 | 82 | 193 (81.4) | 225 (94.9) | 1 | <0.0001 | <0.0001 |
| **Advanced Questions (3<sup>rd</sup> quartile)** |  |  |  |  |  |  |  |
| Higher | 247 | 69.6 | 134 (54.3) | 193 (78.1) | 0.005 | 0.32 | <0.0001 |
| Lower | 207 | 69.9 | 145 (70) | 188 (90.8) | 1 | <0.0001 | <0.0001 |
| **Difficult Questions (4<sup>th</sup> quartile)** |  |  |  |  |  |  |  |
| Higher | 262 | 48.5 | 110 (42) | 153 (58.4) | 1 | 0.23 | 0.002 |
| Lower | 223 | 48.5 | 102 (45.7) | 163 (73.1) | 1 | <0.0001 | <0.0001 |

Chi-squared test was used to calculate p-values. P-values were adjusted for multiple testing using the Bonferroni correction.

### Performance by Topic

A comparative evaluation of GPT-3.5, GPT-4, and the average user performance across various topics was carried out (as depicted in Figure 1). In the “Behavioral, Cognitive, Psych” category, GPT-4 outperformed both GPT-3.5 and average test bank users (GPT-4: 433/482 (89.8%), GPT-3.5: 362/482 (75.1%), human users: 76%, p<0.0001). GPT-4 also exhibited superior performance in topics such as Basic Neuroscience, Movement Disorders, Neurotoxicology, Nutrition, Metabolic, Oncology, and Pain, when compared to GPT-3.5, whereas its performance aligned with the human user average (Table 1, Figure 1).

**Figure 1:**
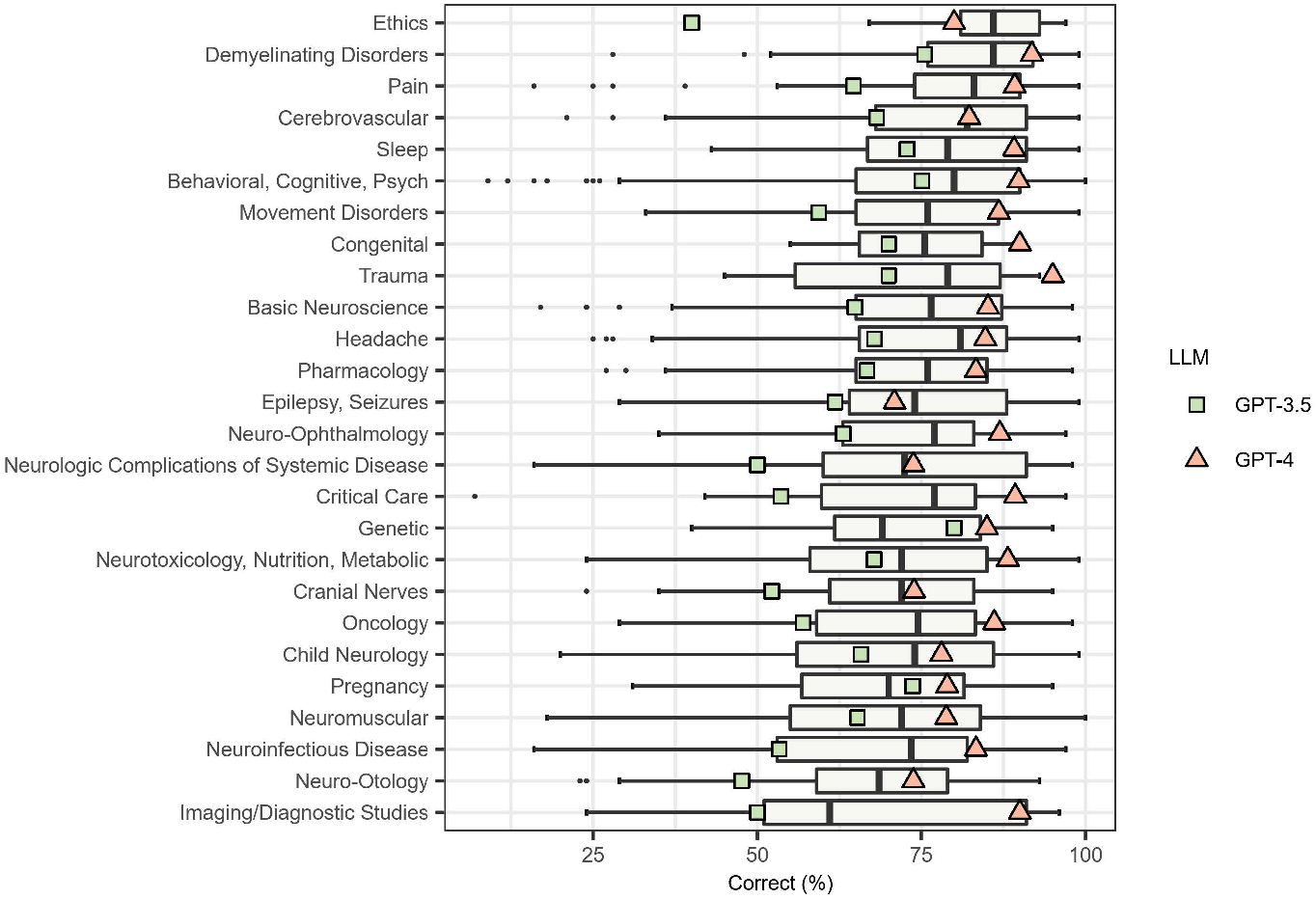
Percentage of correctly answered questions per topic. Human user score distribution is shown by boxplots (grey), LLM performance is shown in green and pink. Percentage of correctly answered questions per topic by human users (grey boxplots, black line indicates median), GPT-3.5 (green) and GPT-4 (pink). In the majority of topics, GPT-4 performs above human average, while GPT-3.5 performs below human average (Table 1).

To identify any topic-specific strengths or weaknesses displayed by each model, we analyzed their performance in topics that contained over 50 questions. Notably, GPT-3.5 did not display any significant performance variation across topics. In contrast, GPT-4 showed significantly enhanced performance in answering questions related to Behavioral, Cognitive and Psych categories (89.8%) compared to its performance on questions concerning Epilepsy, Seizures (39/55 (70.9%), p=0.008) and Neuromuscular topics (145/184 (78.8%), p=0.02).

### Level of Confidence

Both GPT-3.5 and GPT-4 consistently responded to multiple-choice questions using confident or highly confident language (100%, 200 of 200 questions, evaluated by investigators). Self-assessment of confidence expressed by GPT-3.5 and GPT-4 in its answers showed a small difference between incorrect and correct responses (mean Likert score, 4.69 vs 4.79; p<0.0001 for GPT-3.5, mean Likert score 4.77 vs. 4.93; p<0.0001, for GPT-4). Incorrect GPT-4 and GPT-3.5 answers were all subjectively assessed by the models as expressing confidence or high confidence (Likert score 4 or 5, GPT-4: 99.3%, 292 out of 294, GPT-3.5: 100%, 650 out of 650, Supplementary Figure 6). When prompted with the correct answer after an incorrect response, GPT-3.5 and GPT-4 responded by apologizing and agreeing with the provided correct answer in all cases (100%, n=100 of 100 incorrectly answered questions, both GPT-3.5 and GPT-4).

### Reproducibility of Responses

Next, we investigated the reproducibility of responses. For this purpose, the same question (n=100) was independently queried 50 times and the percentage of each question was recorded. Next, we compared answers with high reproducibility (defined as more than 75% of all queries answered with the same answer) to answer without high reproducibility. This analysis revealed that highly reproducible answers are more likely to be answered correctly (66 of 88 (75%)) than inconsistent answers, (5 of 13 (38.5%), p=0.02) by GPT-3.5, potentially indicating another marker of confidence of LLMs that might be leveraged to filter out invalid responses. The same observation was made with GPT-4 with 78 of 96 (81.3%) correct answers in answers with high reproducibility compared to 1 of 4 (25%) in answers with low reproducibility (p=0.04).

### Characteristics of questions using high-dimensional representation analysis of question embeddings

We identified an association of question word length and the ability to answer questions correctly in GPT-

3.5 and GPT-4, incorrectly answered questions being longer on average (Supplementary Figure 7). This was not found in human users, but instead a weak positive correlation between question length and correct answers was observed (R=0.074, p=0.001, Supplementary Figure 7).

When analyzing the high-dimensional representation of correctly and incorrectly answered questions, no pattern into distinct clusters was observed (Supplementary Figure 8).

To investigate whether the models use the similarity of question and answers in the multidimensional embedding space to select their answer, similarity between the question embedding and each answer embeddings was compared. It was found that in 28.3 % of questions (476 of 1681), the correct answer was the closest in the multidimensional embedding space. Accordingly, the LLMs labeled the most similar answer as correct in 30.5% of cases (GPT-4, 513 of 1681, p=0.17, GPT-3.5, 524 of 1681 (31.1%), p=0.08), indicating that the distance between question and answer did not significantly affect the models’ answer choice.

### Qualitative evaluation of GPT-3.5 and GPT-4

All investigators felt that both the performance of GPT-3.5 and especially GPT-4 was impressive. While the performance of GPT-3.5 was sometimes variable and difficult to predict this was more stable with GPT-4. While GPT-3.5 was able to answer some challenging questions correctly, it also incorrectly answered some questions the reviewers perceived as simple. This was not the case with GPT-4 where mostly questions that were perceived as difficult were incorrectly answered. Interestingly, a small portion of incorrect answers was due to flawed reasoning potentially indicating problems of LLMs without any underlying world model ^22^.

## Discussion

The notable progress achieved by GPT-3.5 and GPT-4 has significantly enhanced the potential of Large Language Models (LLMs) across a wide range of applications ^23-25^. Despite being extensively pretrained on vast data sets, offering promising possibilities within the healthcare sector, their specific application in neurology remains relatively uncharted territory. The efficacy of these models in handling specialized neurology knowledge also remained indeterminate until this study.

This exploratory research revealed GPT-4’s proficiency in completing a neurology board-like examination, a task GPT-3.5 was unable to accomplish. This finding underscores the rapid and significant evolution of LLMs.

Despite their strengths, GPT-3.5 and GPT-4 demonstrated subpar performance in tasks requiring higher-order thinking ^13^. Yet, GPT-4 still managed to perform satisfactorily. In comparison to its performance on the USMLE Step Examinations ^6,26,27^, where it did not exceed a 65% accuracy, GPT-3.5 scored surprisingly well in this more specialized examination.

As these models are trained to identify patterns and relationships among words in their training data, they can struggle in situations requiring a deeper understanding of context or specialized technical language. This limitation is crucial for neurologists to bear in mind, particularly with LLMs now being incorporated into popular search engines and readily accessible to the public ^28^.

Interestingly, both models exhibited confident language when answering questions, even when their responses were incorrect. This trait is a recognized limitation of LLMs ^29,30^ and originates from the training objective of these models, which is to predict the most likely sequence of words following an input. This characteristic, coupled with the model’s inclination to generate plausible, convincing, and human-like responses, can potentially mislead individuals relying solely on it for information ^31,32^. However, the model was able to partially differentiate its own confidence level as there were slight but significant differences between correctly and incorrectly answers although the values on the Likert scale predominantly are between “confident” and “highly confident”. Furthermore, we identified that reproducible answers are correlated with correctness and might serve as an intrinsic, surrogate marker of confidence defined by the output of the LLM.

This study has some limitations. The questions used were mostly not official ABPN or EBN questions due to their confidential and regulated nature. Additionally, image-based questions were not included as GPT-3.5 and current versions of GPT-4 are not equipped to process these. Furthermore, the passing grade was an approximation based on the threshold by the ABPN for approving of points for certified medical education. The limited number of questions in each subgroup in this exploratory study also reduced the power of subgroup analyses. We only included GPT-3.5 and GPT-4 in this assessment as other similarly powerful closed-source models were not available to us at the time of this study. Lastly, qualitative assessments of GPT-3.5 and GPT-4 responses are inherently subjective. In conclusion, this study underscored the vast potential of LLMs such as GPT-4, particularly in neurology, even without neurology-specific pretraining. GPT-4 passed a neurology board-style examination after exclusion of video and image questions. As deep learning architectures are continuously refined for computer vision and medical imaging ^33,34^, this image-processing limitation may be addressed in future AI models, potentially including the upcoming multimodal input functionalities of GPT-4. Despite performing admirably on questions assessing basic knowledge and understanding, the model showed slightly lower performance on higher-order thinking questions. Consequently, neurologists should be aware of these limitations, including the models’ tendency to phrase inaccurate responses confidently, and should be cautious regarding its usage in practice or education. With the anticipated advancements in LLMs, neurologists and experts of other clinical disciplines will need to comprehend their performance, reliability, and applications within neurology better. Investigating the potential applications of LLMs that have been fine-tuned specifically for neurology represents a compelling direction for future research.

