## Supplementary Material for "Evaluating the Performance of Large Language Models on a Neurology Board-Style Examination"

**Supplementary Table 1: Performance of GPT-3.5 and GPT-4 on the EBN question samples cohort**

| Question Type | Questions N | GPT-3·5 Correct N (%) | GPT-4 Correct N (%) | Adj P Value GPT-3·5 vs GPT-4 |
|---|---|---|---|---|
| **All Questions** | 19 | 10 (52·6) | 14 (73·7) | 0·31 |
| **Order of thinking** | | | | |
| **Higher** | 14 | 7 (50) | 11 (78·6) | 0·47 |
| **Lower** | 5 | 3 (60) | 3 (60) | 1 |

Chi-squared test was used to calculate p-values. P-values were adjusted for multiple testing using the *Bonferroni* correction.

**Supplementary Figure 1: Example of a correctly, answered lower-order question and answers by GPT-3.5 and GPT-4**

| **Figure 1: Example of a lower-order question, answered correctly** | |
|---|---|
| Question | Explanation by GPT-4 |
| What kind of identification inability does prosopagnosia reflect?<br><br>A: Facial expression.<br><u>B: Familiar faces.</u><br>C: Melody in language.<br>D: Non-musical sounds.<br>E: Pitch of a melody. | B: Familiar faces<br>Prosopagnosia, also known as face blindness, is a cognitive disorder that affects the ability to recognize familiar faces, including one's own face (self-recognition). It does not affect a person's ability to recognize and interpret facial expressions, sounds, or melodies. It is typically associated with damage to the right fusiform gyrus, a region of the brain that is believed to be critical for face perception. |

**Supplementary Figure 2: Example of a correctly, answered higher-order question and answers by GPT-3.5 and GPT-4**

| **Figure 2: Example of a higher-order question, answered correctly** | |
|---|---|
| Question | Explanation by GPT-4 |
| A 51-year-old male, with a history of herpetic shingles in dermatomes T7-10 three years ago complains about pain in the back side of his right leg. He mentioned that there has been some red discoloration of the skin on his right lower leg, but definitely no shingles. On physical examination, he demonstrated normal tone and full strength in all muscle groups bilaterally; reflexes were normal apart from the knee-reflex being somewhat decreased on the right; the adductor reflexes were symmetric. MRI of the lumbal spine showed degenerative disc disease at all lumbar levels, with slight bilateral foraminal stenosis at L3 and L4, but no herniated disc. Results of an electromyogram were normal. His cerebrospinal fluid showed a predominantly lymphocytic pleocytosis (110 cells/L), a raised protein level (570mg/L) and intrathecal Ig-synthesis. Which diagnosis most likely explains the clinical features of this patient?<br><br><u>A: Neuroborreliosis.</u><br>B: Neurosarcoidosis.<br>C: Inflammatory plexopathy.<br>D: Postherpetic neuralgia.<br>E: Wartenberg syndrome. | A: Neuroborreliosis.<br>Neuroborreliosis is a complication of Lyme disease (caused by the bacterium Borrelia burgdorferi), where the bacteria infect the nervous system. The features of this case that suggest neuroborreliosis include the pain in the right leg, the lymphocytic pleocytosis and raised protein level in the cerebrospinal fluid, and the evidence of intrathecal Ig-synthesis, which indicates an immune response within the central nervous system.<br>Postherpetic neuralgia is a complication of shingles and would typically present with pain in the area of the previous shingles rash, which does not appear to be the case here. Neurosarcoidosis, inflammatory plexopathy, and Wartenberg syndrome would likely present with different clinical and diagnostic findings. It's worth noting that Lyme disease is often associated with a characteristic "bull's-eye" rash (erythema migrans), but it may not appear in all cases, and the disease can still progress to neuroborreliosis without it. Further diagnostic testing, such as serological tests for antibodies against Borrelia burgdorferi, would be needed to confirm the diagnosis. |

**Supplementary Figure 3: Example of an incorrectly, answered lower-order question and answers by GPT-3.5 and GPT-4**

| **Figure 3: Example of a lower-order question, answered incorrectly** | |
|---|---|
| Question | Explanation by GPT-4 |
| Which of the following symptoms is found with increased frequency in patients with neuropathic pain as compared with patients with nociceptive pain?<br><br>A: Deep aching pain.<br>B: Lancinating pain.<br>C: Pain evoked by local pressure.<br>D: Pain on exercise.<br>E: Sensation of pins and needles. | B: Lancinating pain.<br>Lancinating pain is a symptom that is found with increased frequency in patients with neuropathic pain as compared with patients with nociceptive pain. This type of pain is characterized by the sudden, sharp, electric shock-like sensation. The other options may also be experienced by patients with neuropathic pain, but they are not specific to this type of pain. |

**Supplementary Figure 4: Example of an incorrectly, answered higher-order question and answers by GPT-3.5 and GPT-4**

| **Figure 4: Example of a higher-order question, correctly** | |
|---|---|
| Question | Explanation by GPT-4 |
| A 35-year-old male was involved in a minor car accident when returning home after a tennis match, and attended hospital because of not being aware of objects on his left side. There was no head injury but nevertheless he felt a throbbing right occipital headache, with no other symptoms. On clinical examination, two hours after the accident, there was a left hemianopia. Eye movements and pupillary function were normal. Blood pressure was 160/95 mmHg. No abnormalities were found on a cranial CT-scan. He reported previous attacks of severe unilateral pulsatile headache with nausea and photophobia between the ages of 15 and 35 years. The frequency of headache was about once in 6 months, with a duration of up to 24 hours. Which diagnosis applies best to this case?<br><br>A: Internal carotid artery stenosis.<br>B: Migrainous infarction.<br>C: Reversible vasoconstriction syndrome.<br>D: Posterior reversible encephalopathy syndrome.<br><u>E: Vertebral artery dissection.</u> | B: Migrainous infarction.<br>The term "migrainous infarction" describes a rare type of ischemic stroke that occurs during a migraine with aura. Given this patient's history of severe, unilateral, pulsatile headaches with nausea and photophobia (typical features of migraines), and the current presentation with headache and visual disturbance (hemianopia), migrainous infarction is a plausible diagnosis.<br><br>The onset of visual symptoms after a minor accident could be coincidental, but it's also possible that the stress of the accident triggered a migraine attack. The normal CT-scan findings don't exclude migrainous infarction, as the changes associated with this condition may not be visible on imaging, especially in the early stages.<br><br>The other options (internal carotid artery stenosis, reversible vasoconstriction syndrome, posterior reversible encephalopathy syndrome, and vertebral artery dissection) could potentially cause similar symptoms, but they don't align as closely with this patient's history and presentation. For example, vertebral artery dissection would typically cause symptoms related to the posterior circulation, such as vertigo, imbalance, or difficulty speaking or swallowing, which this patient doesn't have. |

**Supplementary Figure 5: Performance of GPT-3.5 and GPT-4 based on difficulty levels, with difficulty being assessed by the percentage of human users who answered correctly**

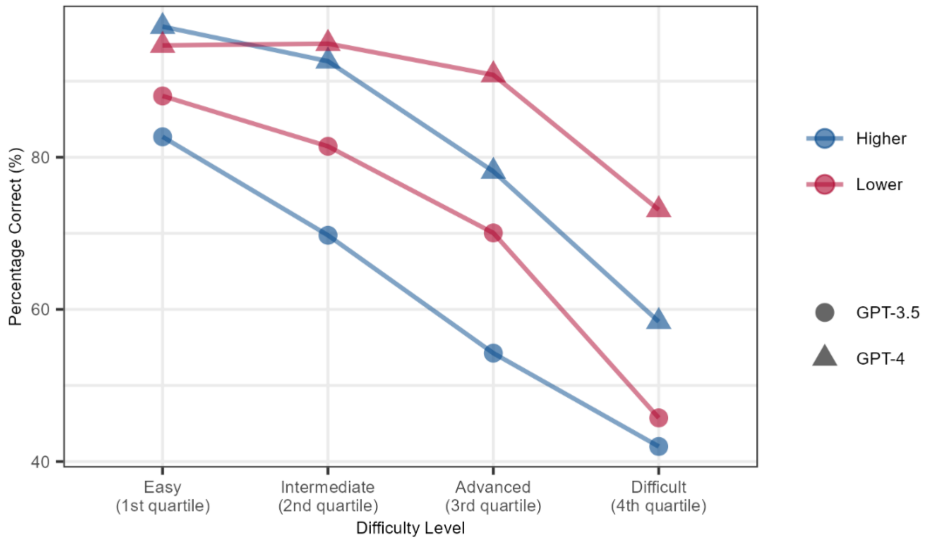

Based on human user performance, questions are categorized into easy: 1st quartile, intermediate: 2nd quartile, advanced: 3rd quartile and difficult: 4th quartile questions. For each group of question, the percentage of correctly answered questions per LLM (symbol) and question type (color) is visualized.

**Supplementary Figure 6: Confidence of language in correctly and incorrectly answered questions**

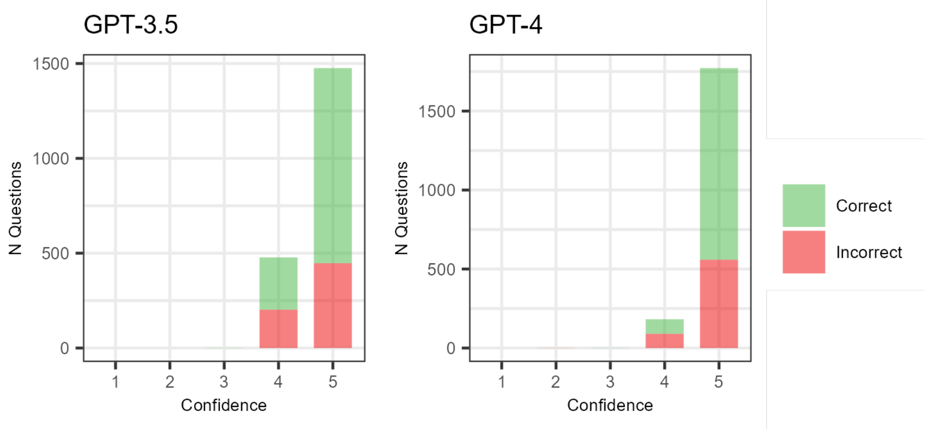

Bar plot visualizing self-assessed confidence on a Likert scale by GPT-3.5 and GPT-4. Questions are colored based on whether they were answered correctly. (N=1956)

**Supplementary Figure 7: Length of question between incorrectly and correctly answered questions between GPT 3.5, GPT 4 and question bank users separately, primary vs high-order question percentage**

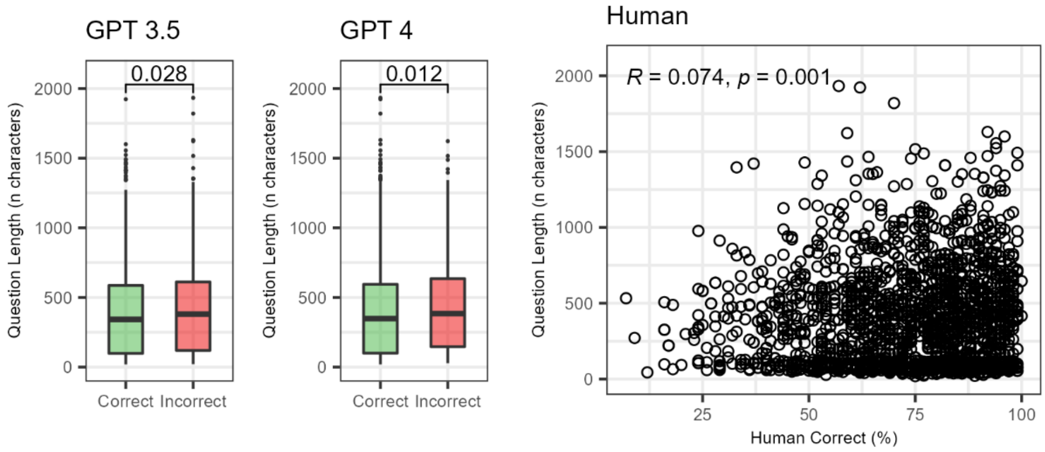

Left: Comparison of Question Length between correctly and incorrectly answered questions for GPT3.5 and GPT4 (N=1956 questions). Right: Correlation plot between the percentage of correctly answering users per question and the question length, one dot representing a single question. (N=1956).

**Supplementary Figure 8: High-dimensional tSNE analyses of question and answer embeddings**

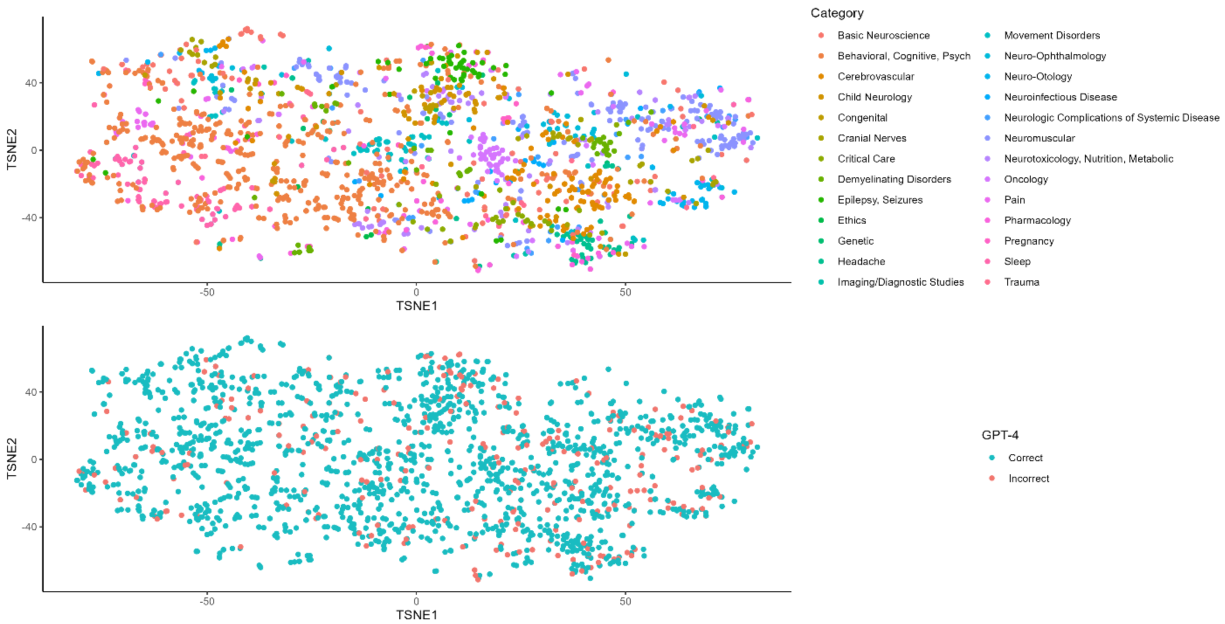

T-SNE analysis of calculated embeddings of questions, each question represented by a single dot and colored based on their related topic.
